# Delta variants of SARS-CoV-2 cause significantly increased vaccine breakthrough COVID-19 cases in Houston, Texas

**DOI:** 10.1101/2021.07.19.21260808

**Authors:** Paul A. Christensen, Randall J. Olsen, S. Wesley Long, Sishir Subedi, James J. Davis, Parsa Hodjat, Debbie R. Walley, Jacob C. Kinskey, Matthew Ojeda Saavedra, Layne Pruitt, Kristina Reppond, Madison N. Shyer, Jessica Cambric, Ryan Gadd, Rashi M. Thakur, Akanksha Batajoo, Regan Mangham, Sindy Pena, Trina Trinh, Prasanti Yerramilli, Marcus Nguyen, Robert Olson, Richard Snehal, Jimmy Gollihar, James M. Musser

## Abstract

Genetic variants of SARS-CoV-2 have repeatedly altered the course of the COVID-19 pandemic. Delta variants of concern are now the focus of intense international attention because they are causing widespread COVID-19 disease globally and are associated with vaccine breakthrough cases. We sequenced the genomes of 16,965 SARS-CoV-2 from samples acquired March 15, 2021 through September 20, 2021 in the Houston Methodist hospital system. This sample represents 91% of all Methodist system COVID-19 patients during the study period. Delta variants increased rapidly from late April onward to cause 99.9% of all COVID-19 cases and spread throughout the Houston metroplex. Compared to all other variants combined, Delta caused a significantly higher rate of vaccine breakthrough cases (23.7% for Delta compared to 6.6% for all other variants combined). Importantly, significantly fewer fully vaccinated individuals required hospitalization. Individuals with vaccine breakthrough cases caused by Delta had a low median PCR cycle threshold (Ct) value (a proxy for high virus load). This value was closely similar to the median Ct value for unvaccinated patients with COVID-19 caused by Delta variants, suggesting that fully vaccinated individuals can transmit SARS-CoV-2 to others. Patients infected with Alpha and Delta variants had several significant differences. Our integrated analysis emphasizes that vaccines used in the United States are highly effective in decreasing severe COVID-19 disease, hospitalizations, and deaths.

## [Introduction]

Delta variants of concern (VOCs) of SARS-CoV-2 such as B.1.617.2, AY.3, and AY.4 are the focus of intense international concern because they are causing widespread COVID-19 disease in the United States, Southeast Asia, Europe, and elsewhere (https://www.cdc.gov/coronavirus/2019-ncov/cases-updates/variant-surveillance/variant-info.html, last accessed: August 18, 2021; https://www.gov.uk/government/collections/new-sars-cov-2-variant, last accessed: August 18, 2021)^1^. For example, Delta has replaced the Alpha variant in the United Kingdom, previously the cause of virtually all COVID-19 cases in that country (https://www.who.int/publications/m/item/weekly-epidemiological-update-on-covid-19---13-july-2021, last accessed August 18, 2021; https://www.ons.gov.uk/peoplepopulationandcommunity/healthandsocialcare/conditions/anddiseases/bulletins/coronaviruscovid19infectionsurveypilot/9july2021, last accessed August 18, 2021). Vaccine breakthrough cases caused by SARS-CoV-2 variants also have become of considerable public health and biomedical concern worldwide^2-6^. To study Delta spread and vaccine breakthrough cases in metropolitan Houston, we sequenced the genomes of 16,965 SARS-CoV-2 from patient samples acquired March 15, 2021 through September 20, 2021 using an Illumina NovaSeq 6000 instrument and methods described previously^7, 8^. This period includes the time from initial identification of Delta-related VOCs in late April in our large Houston Methodist healthcare system until Delta VOCs continuously caused the supermajority (99.9%) of all new cases. The sequenced sample is 91% of the 18,736 total COVID-19 cases diagnosed in our health system during the study period. We also compared characteristics of patients infected with Alpha and Delta VOCs, an analysis that identified several significant differences between patients infected with these two variants.

## Materials and Methods

### Patient Specimens

Specimens were obtained from registered patients at Houston Methodist hospitals, associated facilities (e.g., urgent care centers), and institutions in the Houston metropolitan region that use our laboratory services. The great majority of individuals had signs or symptoms consistent with COVID-19 disease. For analyses focusing on the Delta family variants, a comprehensive sample of genomes obtained from March 15, 2021 through September 20, 2021 was used. This time frame was chosen because it represents the period during which the downturn of our third wave was occurring, and soon after we identified the first Delta variant case in our health care system.

Subsequently, Delta variants increased rapidly to continuously cause 99.9% of all new cases. The study included 16,965 unique patients identified in this time frame for whom we had SARS-CoV-2 genome sequences. For analyses comparing features of patients infected with the Delta variants and Alpha VOC, a comprehensive sample of genomes obtained from January 1, 2021 through September 20, 2021 was used. This time frame represents the period during which we identified the first Alpha variant case in our health care system. This VOC increased rapidly and peaked, and then decreased to cause less than 1% of all cases in the region. This part of the study included 28,560 unique patients identified in the January 1, 2021 through September 20, 2021 period. The work was approved by the Houston Methodist Research Institute Institutional Review Board (IRB1010-0199).

### SARS-CoV-2 Molecular Diagnostic Testing

Specimens obtained from symptomatic patients with a suspicion for COVID-19 disease were tested in the Molecular Diagnostics Laboratory at Houston Methodist Hospital using assays granted Emergency Use Authorization (EUA) from the FDA (https://www.fda.gov/medical-devices/emergency-situations-medical-devices/faqs-diagnostic-testing-sars-cov-2#offeringtests, last accessed June 7, 2021). Multiple molecular testing platforms were used, including the COVID-19 test or RP2.1 test with BioFire Film Array instruments, the Xpert Xpress SARS-CoV-2 test using Cepheid GeneXpert Infinity or Cepheid GeneXpert Xpress IV instruments, the cobas SARS-CoV-2 & Influenza A/B Assay using the Roche Liat system, the SARS-CoV-2 Assay using the Hologic Panther instrument, the Aptima SARS-CoV-2 Assay using the Hologic Panther Fusion system, the Cobas SARS-CoV-2 test using the Roche 6800 system, and the SARS-CoV-2 assay using Abbott Alinity m instruments. Virtually all tests were performed on material obtained from nasopharyngeal swabs immersed in universal transport media (UTM); oropharyngeal or nasal swabs, bronchoalveolar lavage fluid, or sputum treated with dithiothreitol (DTT) were sometimes used. Standardized specimen collection methods were used (https://vimeo.com/396996468/2228335d56, last accessed June 7, 2021).

### SARS-CoV-2 Genome Sequencing

Libraries for whole SARS-CoV-2 genome sequencing were prepared according to version 3 or version 4 (https://community.artic.network/t/sars-cov-2-version-4-scheme-release/312, last accessed August 19, 2021) of the ARTIC nCoV-2019 sequencing protocol. Our semi-automated workflow described previously^7, 8^ employed BioMek i7 liquid handling workstations (Beckman Coulter Life Sciences) and MANTIS automated liquid handlers (FORMULATRIX). Short sequence reads were generated with a NovaSeq 6000 instrument (Illumina). For continuity of the epidemiologic analysis in the study period, we included some genome sequences reported in a recent publication.^8^

### SARS-CoV-2 Genome Sequence Analysis and Identification of Variants

Viral genomes were assembled with the BV-BRC SARS-Cov2 assembly service (https://www.bv-brc.org/app/ComprehensiveSARS2Analysis, last accessed June 7, 2021, requires registration). The One Codex SARS-CoV-2 variant calling and consensus assembly pipeline was used to assemble all sequences (https://github.com/onecodex/sars-cov-2.git, last accessed June 7, 2021) using default parameters and a minimum read depth of 3. Briefly, the pipeline uses seqtk version 1.3-r116 for sequence trimming (https://github.com/lh3/seqtk.git, last accessed June 7, 2021); minimap version 2.1 for aligning reads against reference genome Wuhan-Hu-1 (NC_045512.2); samtools version 1.11 for sequence and file manipulation; and iVar version 1.2.2 for primer trimming and variant calling. Genetic lineages, VOCs, and variants of interest (VOIs) were identified based on genome sequence data and designated by Pangolin v. 3.1.11 with pangoLEARN module 2021-08-09 (https://cov-lineages.org/resources/pangolin.html, last accessed August 18, 2021).

There is a known problem with the ARTIC V3 primer set that results in lack of detection or ambiguity of the G142D and D950N single nucleotide variants, even though these amino acid changes (142D and 950N) are present in the overwhelming majority of B.1.617.2 Delta variant isolates (https://community.artic.network/t/sars-cov-2-version-4-scheme-release/312, last accessed September 1, 2021, https://outbreak.info/compare-lineages?pango=B.1.617.2&gene=S&threshold=0.2, last accessed September 1, 2021). After we transitioned to ARTIC V4, we observed that all Delta variant strain haplotypes contained both 142D and 950N polymorphisms except for four isolates, two of which lacked 142D and two which lacked 950N. We recently reported an analysis that confirmed the previously identified ARTIC V3 problem^9^. Thus, for the Delta isolates presented here, virtually all samples sequenced with ARTIC V3 are presumed to have G142D and D950N.

### Patient Metadata and Geospatial Analysis

Patient metadata (**Table 1, Table 2, Table 3**) were acquired from the electronic medical record by standard informatics methods. Patient home address zip codes were used to visualize the geospatial distribution of spread for each VOC and VOI. Figures were generated with Tableau version 2020.3.4 (<u>Tableau Software, LLC, Seattle, WA</u>). A vaccination breakthrough case was defined as a PCR-positive sample from a patient obtained greater than 14 days after full vaccination (i.e., both doses of the Pfizer or Moderna mRNA vaccines) was completed. For some cases, manual chart review was conducted to resolve discrepancies or ambiguities.

**Table 1.**
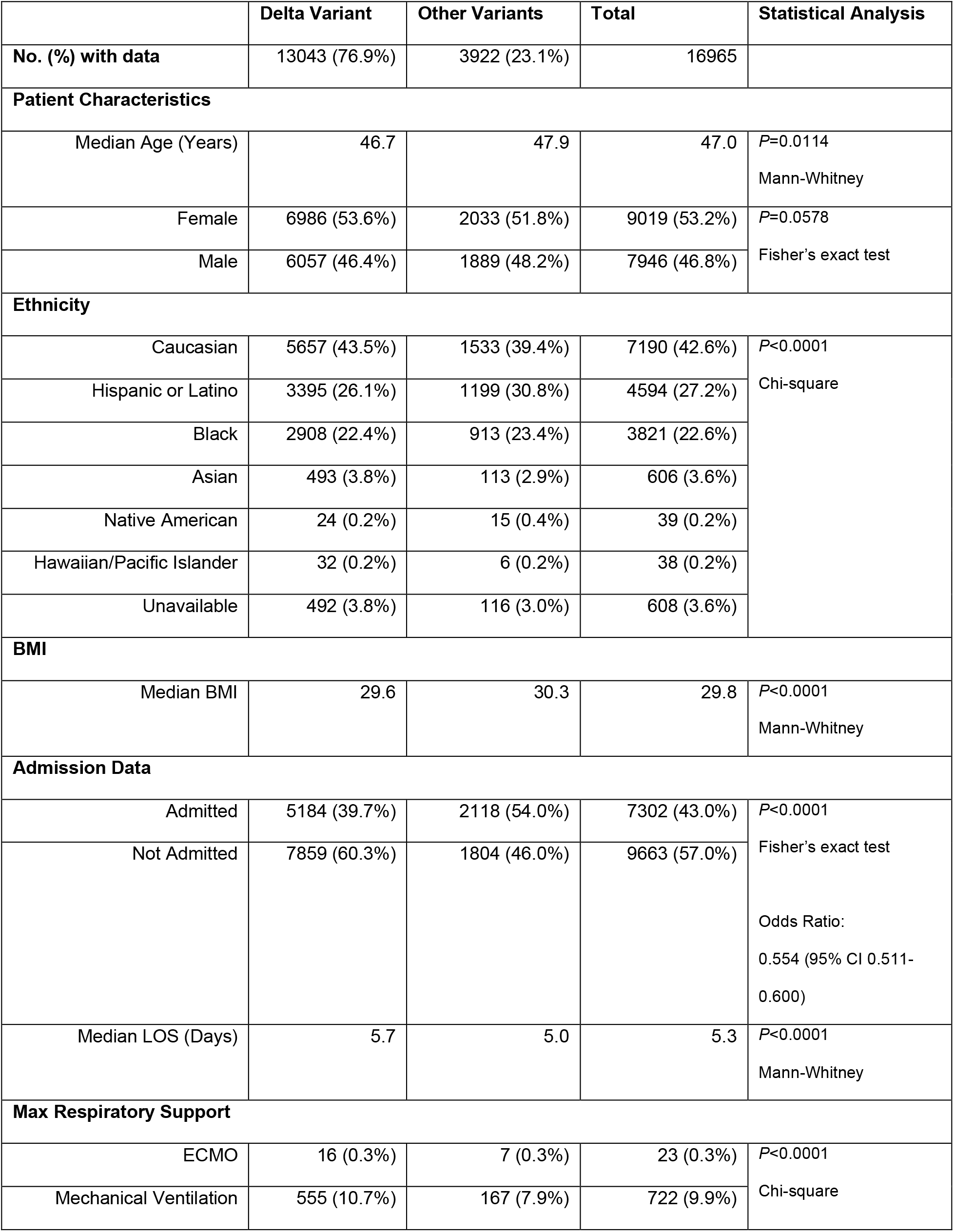
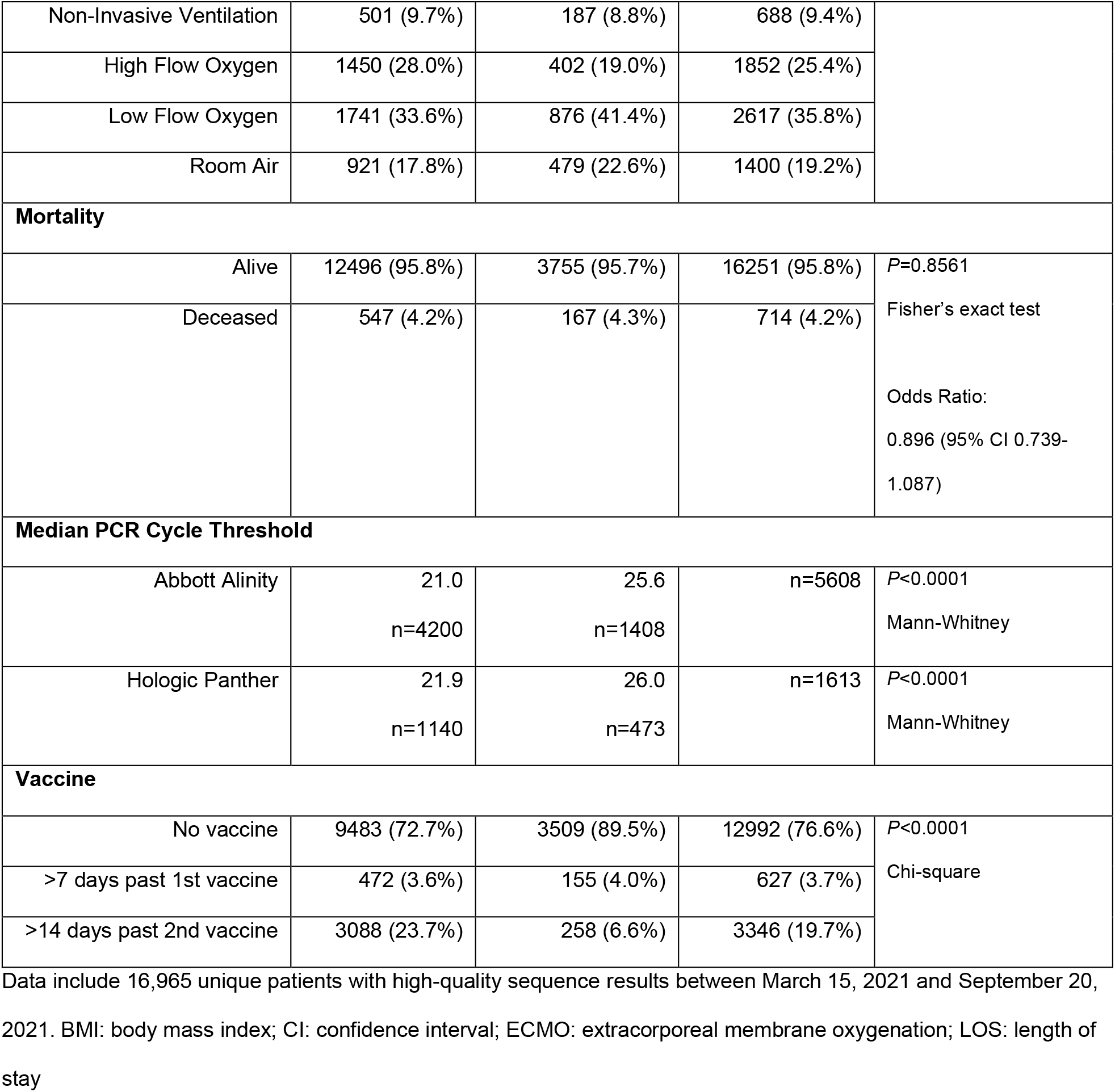
Summary of pertinent patient metadata for the 16,965 unique patients.

**Table 2.**
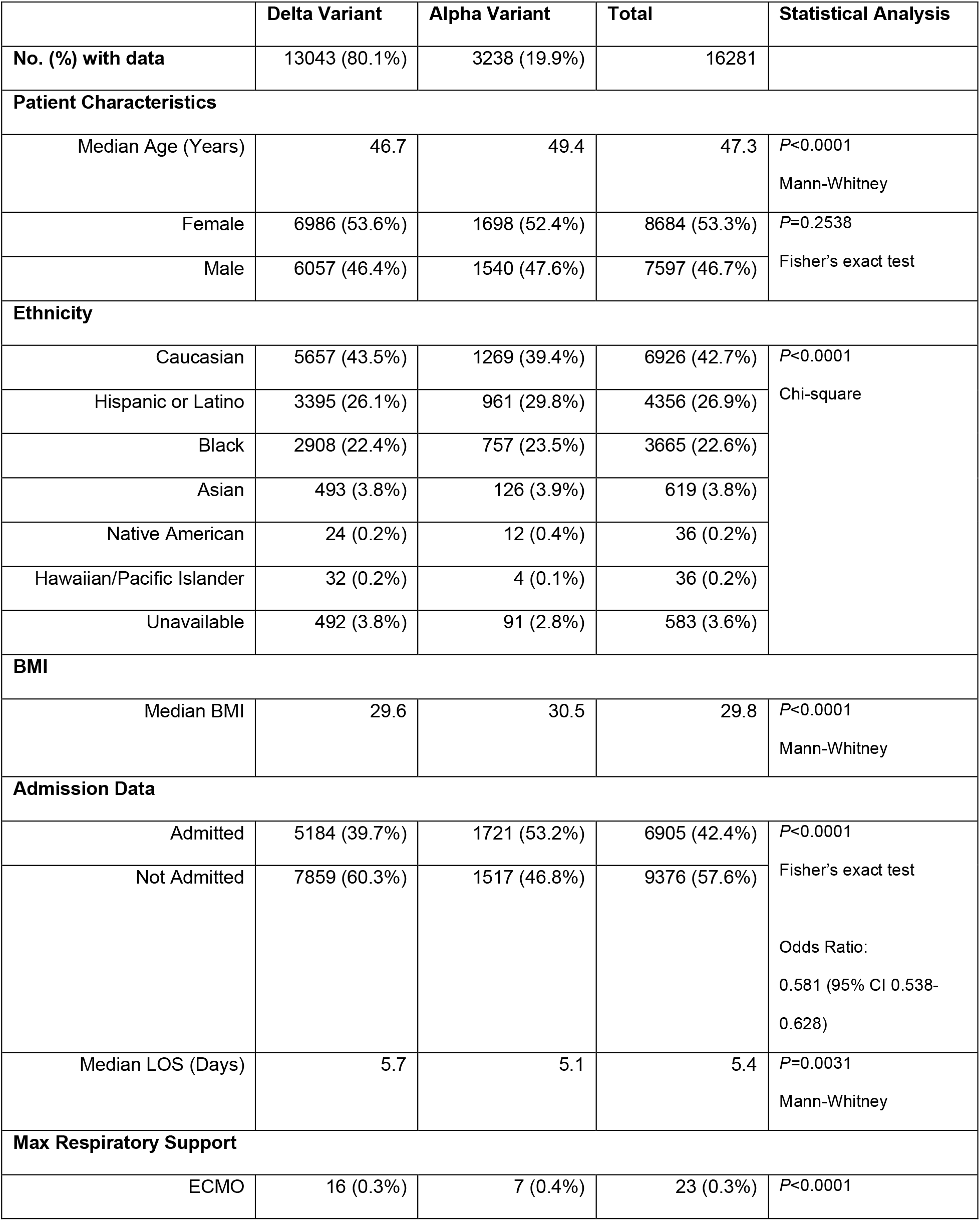
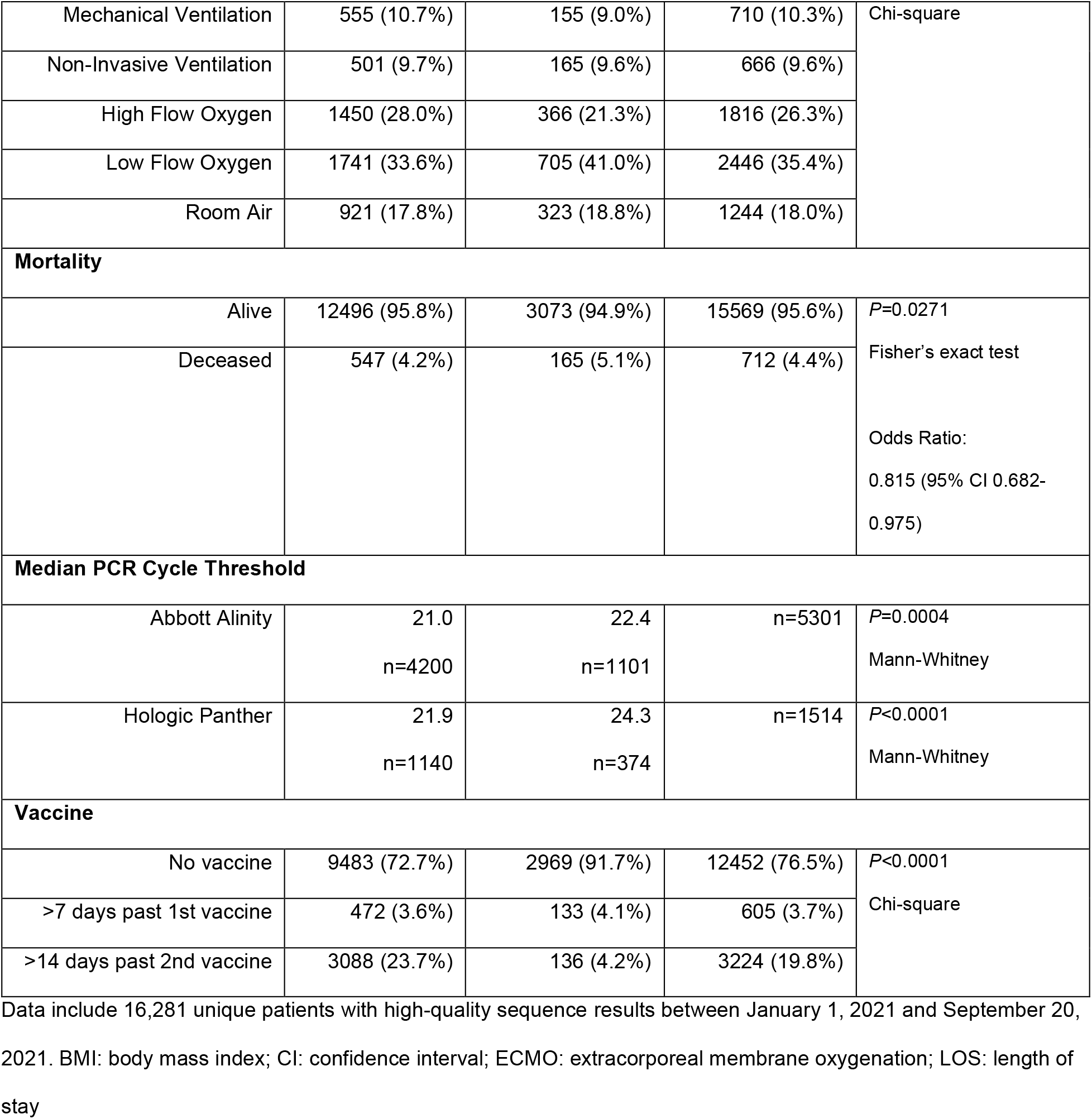
Comparison of pertinent patient metadata for 16,281 unique Delta VOC and Alpha VOC patients.

**Table 3.** Summary of pertinent patient metadata for the 3,346 fully vaccinated patients.

|  | Vaccinated | Not Vaccinated | Total | Statistical Analysis |
| --- | --- | --- | --- | --- |
| Median PCR Cycle Threshold |  |  |  |  |
| Abbott Alinity | 20.5<br>n=1244 | 22.1<br>n=4364 | n=5608 | P=0.0018<br>Mann-Whitney |
| Hologic Panther | 22.2<br>n=378 | 23.5<br>n=1235 | n=1613 | P=0.0348<br>Mann-Whitney |
| Admission Data |  |  |  |  |
| Admitted | 896 (26.8%) | 6406 (47.0%) | 7302 (43.0%) | P<0.0001<br>Fisher's exact test<br>Odds Ratio:<br>0.412 (95% CI 0.379-0.448) |
| Not Admitted | 2450 (73.2%) | 7213 (53.0%) | 9663 (57.0%) |  |
|  | Delta Variant | Other Variants | Total |  |
| No. (%) with data | 3088 (92.3%) | 258 (7.7%) | 3346 |  |
| Admission Data |  |  |  |  |
| Admitted | 800 (25.9%) | 96 (37.2%) | 896 (26.8%) | P=0.0001 |
| Not Admitted | 2288 (74.1%) | 162 (62.8%) | 2450 (73.2%) | Fisher's exact test<br>Odds Ratio:<br>0.590 (95% CI 0.453-0.769) |

## Results

### Delta Epidemiologic Wave

The first Houston Methodist patient infected with a Delta family variant was identified in mid-April 2021, a time when the Alpha VOC was responsible for most COVID-19 cases in metropolitan Houston, and the area was experiencing a steady downturn in total number of new COVID-19 cases (**Figure 1**). Delta VOCs slowly increased in frequency, but beginning in early July a sharp uptick of COVID-19 cases caused by these VOCs occurred (**Figure 1, Figure 2**), with an estimated doubling time of approximately seven days. By September 20, the genome data showed that Delta VOCs accounted for 99.9% of all new COVID-19 cases in our health system (**Figure 2**). This represents the fourth wave of COVID-19 cases in metropolitan Houston (**Figure 1**).

**Figure 1.**
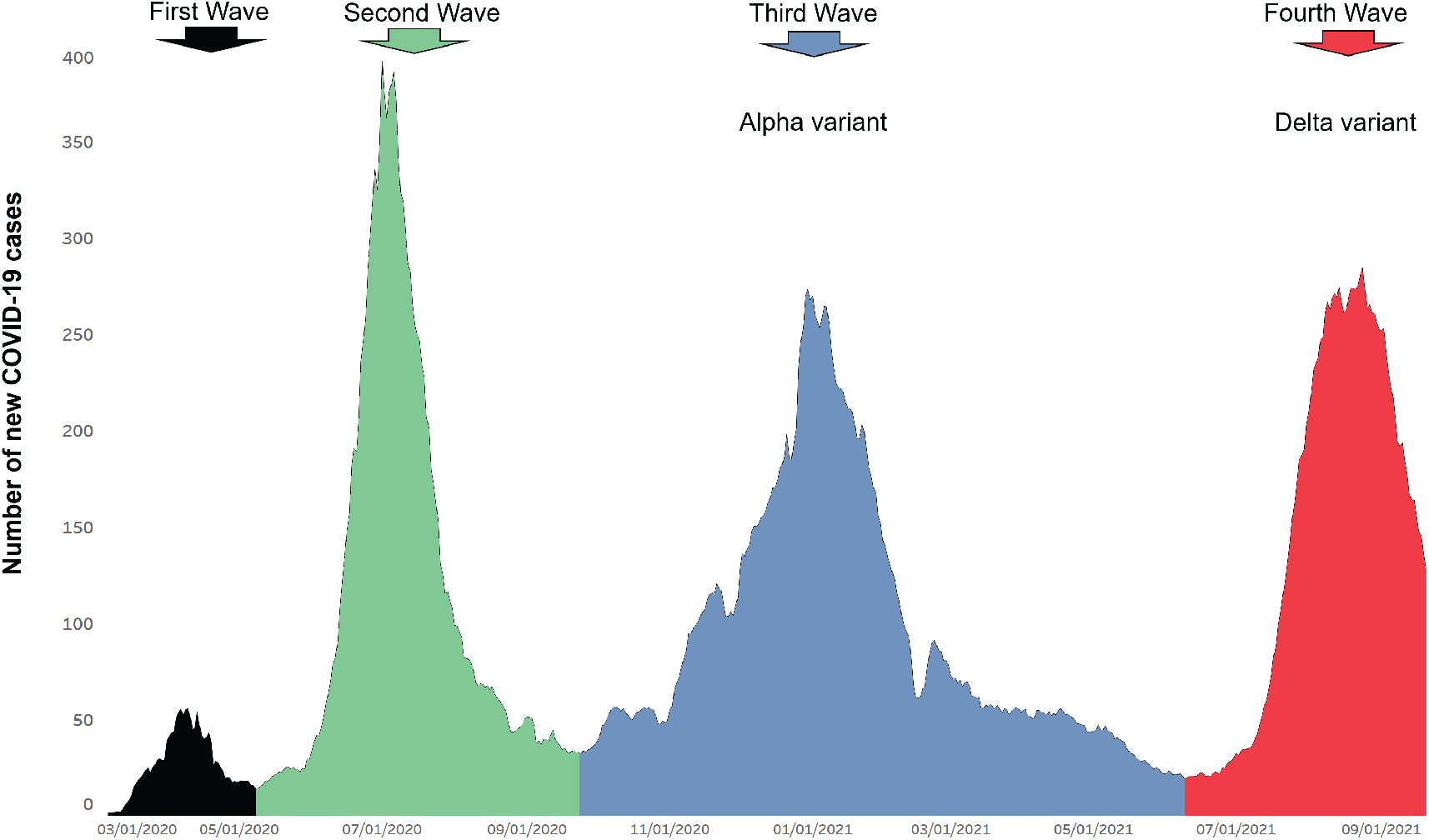
Epidemiologic curve showing four COVID-19 disease waves in Houston Methodist patients. Number of new COVID-19 cases (y-axis) totals are shown as a +/-three-day moving average. Each of the four waves is shown in a different color simply as a stylistic device. The first and second waves were composed of a heterogenous array of SARS-CoV-2 genotypes. The Alpha variant shown in the third wave and the Delta variant shown in the fourth indicate their numeric prominence in those waves, and should not be interpreted to mean that all cases in the third and fourth waves were caused by Alpha and Delta VOCs, respectively. The fourth wave shown includes data through September 20, 2021. The figure was generated with Tableau version 2020.3.4 (Tableau Software, LLC, Seattle, WA). The curve mirrors that of disease activity in metropolitan Houston, Texas.

**Figure 2.**
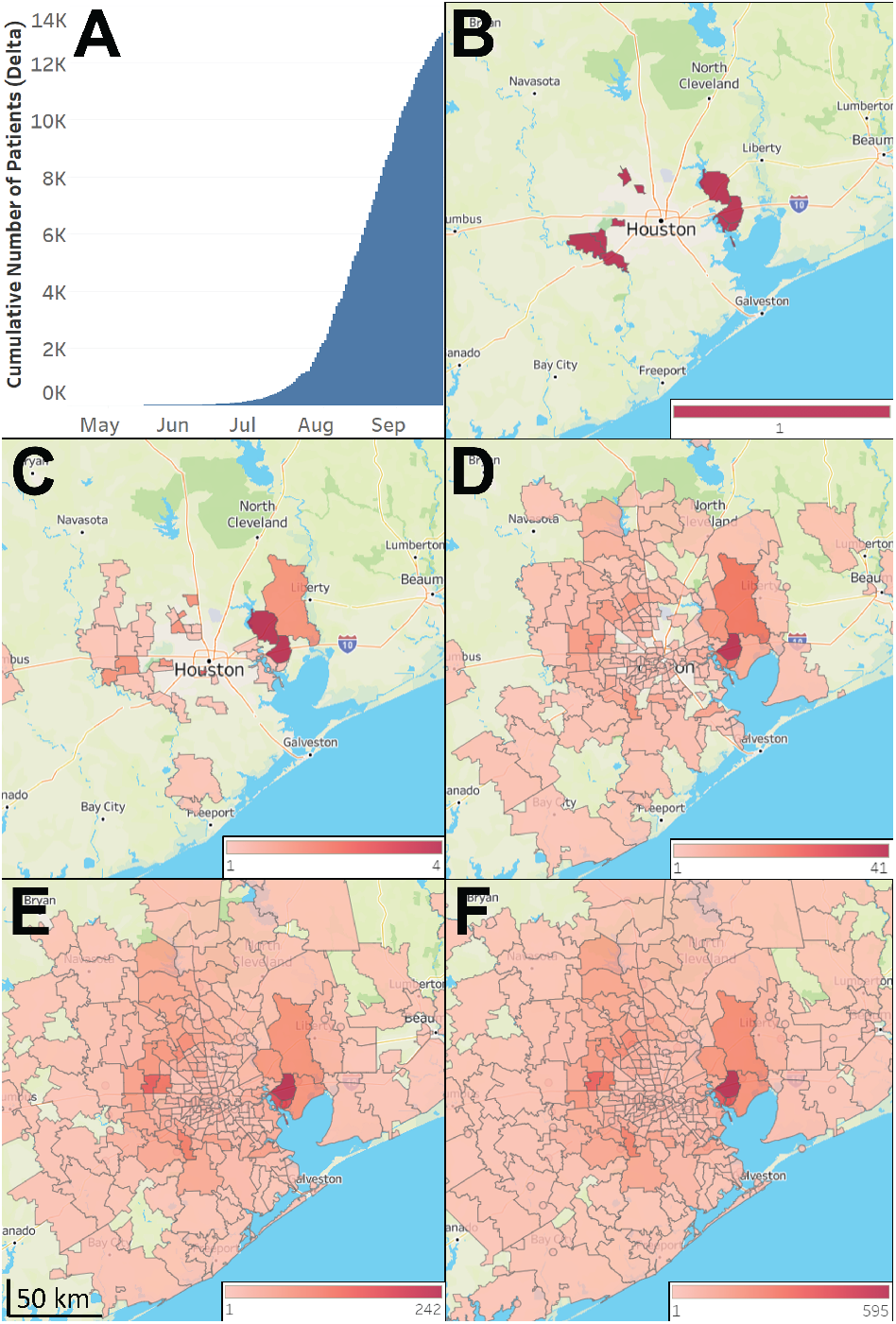
Increase in Delta frequency over time and distribution in metropolitan Houston. The study time frame was March 15, 2021 through September 20, 2021. **A:** Cumulative increase in Delta during the study period; y-axis is the cumulative number of new COVID-19 cases. At the end of the study period, Delta continuously caused 99.9% of all COVID-19 cases. **B – E:** Geospatial distribution of Delta based on home address zip code for each patient. **B**: April 15 – May 15; **C**: April 15 – June 15; **D**: April 15 – July 15; **E**: April 15 – August 15; **F**: April 15 – September 20. Note differences in heat map scale for each panel. Figures were generated using Tableau version 2020.3.4. (Tableau Software, LLC, Seattle, WA).

During the study period 18,736 COVID-19 cases were diagnosed in our healthcare system, and we sequenced 16,965 SARS-CoV-2 genomes, representing 91% of cases. We identified 13,043 patients (76.9% of the total sequenced) with Delta VOCs (Table 1A). The majority (62%; 2,418 of 3,922) of the non-Delta COVID-19 cases occurring during the study period were caused by the Alpha (UK B.1.1.7) VOC.

Consistent with extensive infections caused by Delta variants in Southeast Asia and elsewhere (https://www.cdc.gov/coronavirus/2019-ncov/cases-updates/variant-surveillance/variant-info.html, last accessed: August 18, 2021; https://www.gov.uk/government/collections/new-sars-cov-2-variant, last accessed: August 18, 2021)^1^, several patients had very recent travel histories to countries with a high prevalence of these VOCs, suggesting acquisition abroad and importation into Houston. Sixty patients with Delta Plus (Delta plus the K417N spike amino acid substitution) were identified.

To understand the geospatial distribution of Delta in metropolitan Houston, we acquired patient metadata from the electronic medical record by standard informatics methods, and home address zip codes were used to visualize virus spread (**Figure 2**). Delta VOCs were widely distributed throughout metropolitan Houston, with 334 distinct zip codes represented (**Figure 2**), indicative of the ability of Delta variants to spread very effectively and rapidly between individuals. Analysis of Delta VOCs dissemination over time illustrated the rapid spread of these variants throughout the Houston metroplex (**Figure 2**).

### Delta Family Subvariants

A total of 1,236 subvariants was identified based on amino acid changes in spike protein. The seven most common identified in our study are shown (**Figure 3**) and these represented 73% of the 13,043 Delta samples. It is possible that the unexpectedly high number of subvariants reflects several contributing factors, including the large population sizes and genetic diversity of Delta globally, and multiple independent introductions into the Houston area. Other factors that in principle may contribute to subvariant generation and survival include high virus load in patients on initial diagnosis (**Table 1**), infection of partially immunized hosts, and prolonged infections occurring in relatively immunocompromised individuals^10-16^. In this regard, some of our patients had a history of cancer and organ transplants (**data not shown**). Patients infected with the same subvariant usually lived in different zip codes and generally had no apparent epidemiological link, consistent with the ability of Delta to spread very rapidly. In some cases there were clear epidemiologic links between patients infected with the same subvariant, including being members of the same household.

**Figure 3.**
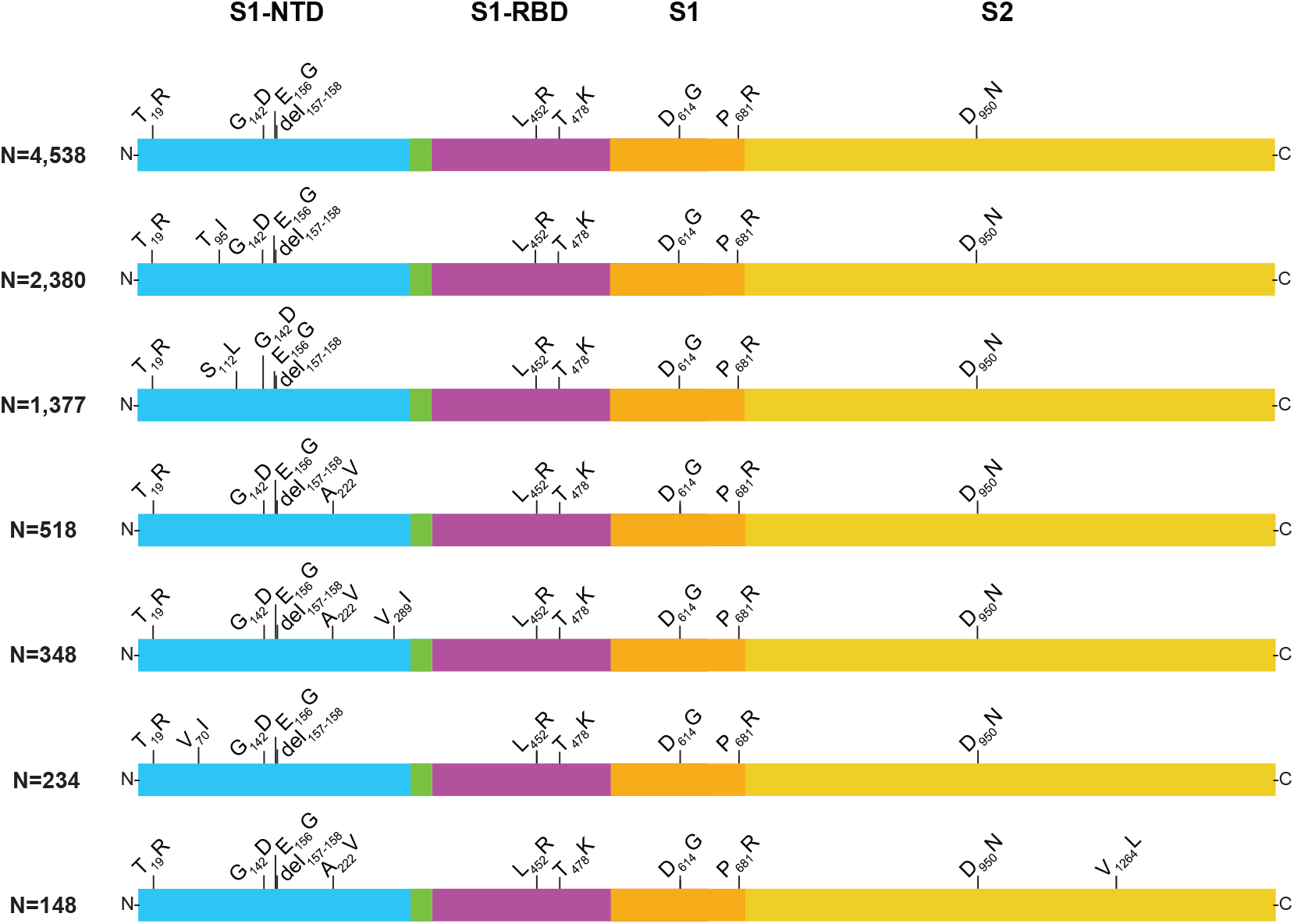
Structural changes present in spike protein of the seven most abundant Delta subvariants identified in the study. The numbers denote the number of patients with each subvariant identified in this study. These seven subvariants represented 73% of the 13,043 Delta samples identified in the study. In this figure, we included all subvariants represented by at least 100 samples. S1-NTD, S1 domain-aminoterminal domain; S1-RBD, S1 domain-receptor binding domain; S1, S1 domain; S2, S2 domain. The figure is a modified version of one published previously^8^.

There is a considerable lack of detailed information about patients in the United States with COVID-19 caused by Delta VOCs (https://www.who.int/publications/m/item/weekly-epidemiological-update-on-covid-19---13-july-2021, last accessed August 18, 2021). Compared to all other patients in the time frame studied, there was no significant difference in sex, but patients infected with Delta were significantly but modestly younger (**Table 1**). However, Delta variant patients were admitted significantly less frequently than other patients and had a significantly longer median hospital length of stay, but there was no significant difference in mortality rate (**Table 1**). Delta cases were more likely to be Asian (consistent with multiple recent entry points from abroad) and Caucasian and less likely to be Hispanic or Latino, have significantly lower PCR cycle threshold (Ct) values on initial diagnosis (a proxy for higher virus load, a finding consistent with increased transmissibility of Delta variants). In addition, Delta variants caused a significantly much higher rate of vaccine breakthrough cases (23.7% compared to 6.6% for all other variants combined) (**Table 1**). Consistent with Delta causing an increased number of vaccine breakthrough cases, it has been reported that this variant has reduced sensitivity to antibody neutralization *in vitro*^14^.

### Comparison of Delta and Alpha COVID-19 Cases

The occurrence of a prominent third wave of COVID-19 cases in Houston caused by the Alpha VOC (**Figure 1**) provided the opportunity of compare and contrast large disease waves caused by two genetically distinct SARS-CoV-2 genotypes. For this analysis we included Alpha VOC cases beginning January 1, 2021, because that time was soon after our first recorded Houston Methodist case caused by this variant. The number of Delta cases studied (n = 13,043) was much greater than the number of Alpha cases (n = 3,238) (**Table 2**). The two patient populations differed significantly in many characteristics, including median age, ethnicity, median PCR Ct level, admission rates, maximum respiratory support, rate of vaccine breakthrough, median length of stay, and mortality (**Table 2**).

### Vaccine Breakthrough Cases

We next analyzed vaccine breakthrough cases (**Table 3**). We found 3,346 of the 16,965 total patients (19.7%) for whom we have genome sequence data met the CDC definition of vaccine breakthrough cases (**Table 1 and Table 3**). There was no simple relationship between the time elapsed since administration of the second booster vaccination and the date of vaccination breakthrough (data not shown). These 3,346 patients received either the Pfizer-BioNTech BNT162b2 (*n* = 2,829, 85%), Moderna mRNA-1273 (*n* = 365, 11%), or J&J/Janssen JNJ-78436735 (*n* = 147, 4%) vaccine; vaccine type was not specified for five individuals. This distribution generally reflects the great majority of BNT162b2 vaccination doses given in our health system and should not be interpreted to mean that the Pfizer-BioNTech and Moderna products are unusually prone to breakthrough cases, as these mRNA vaccines are highly efficacious^6, 17-23^. Vaccinated patients infected with non-Delta variants had a significantly higher Ct value on initial diagnosis, likely indicating better vaccine protection, lower virus load, and decreased probability of transmission. Compared to all other variants combined, a significantly lower percentage (25.9% compared to 37.2%; *P*=0.0001) of patients with breakthrough cases caused by Delta variants was admitted to the hospital (**Table 1 and Table 3**).

### Lambda and Mu Variants of Interest

The Lambda variant of interest was first reported in November 2020. It has very recently attracted widespread media and public interest because it has caused large numbers of infections in Peru and Ecuador, and has been reported to cause COVID-19 in the U.S^24,25^ (https://www.who.int/en/activities/tracking-SARS-CoV-2-variants/, last accessed September 2, 2021). There has been speculation that Lambda may become numerically prominent in the United States and other countries, in part because its spike protein differs immunologically and has been reported to be partially resistant *in vitro* to vaccine-induced sera and therapeutic monoclonal antibodies^26-28^. Because of this concern, we inspected our large genome sequence database to determine if Lambda was present, and if so, had it increased substantially in the Houston metropolitan region. The analysis identified only eight cases caused by this variant of interest, the first one identified in our Houston Methodist patient population in mid-July 2021 (**Figure 4A**). These patients lived in several non-contiguous zip codes throughout the metroplex, that is, were not restricted to a single geographic area of greater Houston (**Figure 4A, B**). The Mu VOI was initially identified in Colombia in January, 2021, and has been reported to account for 39% of cases in that country^29^. Mu also has been found in many other countries worldwide, and recently it was reported that the Mu variant has increased resistance *in vitro* to antibodies elicited by vaccination with BNT162b2 and natural infection by SARS-CoV-2^30^. We identified 53 cases of COVID-19 caused by the Mu VOI, and similar to the Lambda these cases were distributed in multiple areas in the Houston metropolitan region (**Figure 4C, D**).

**Figure 4.**
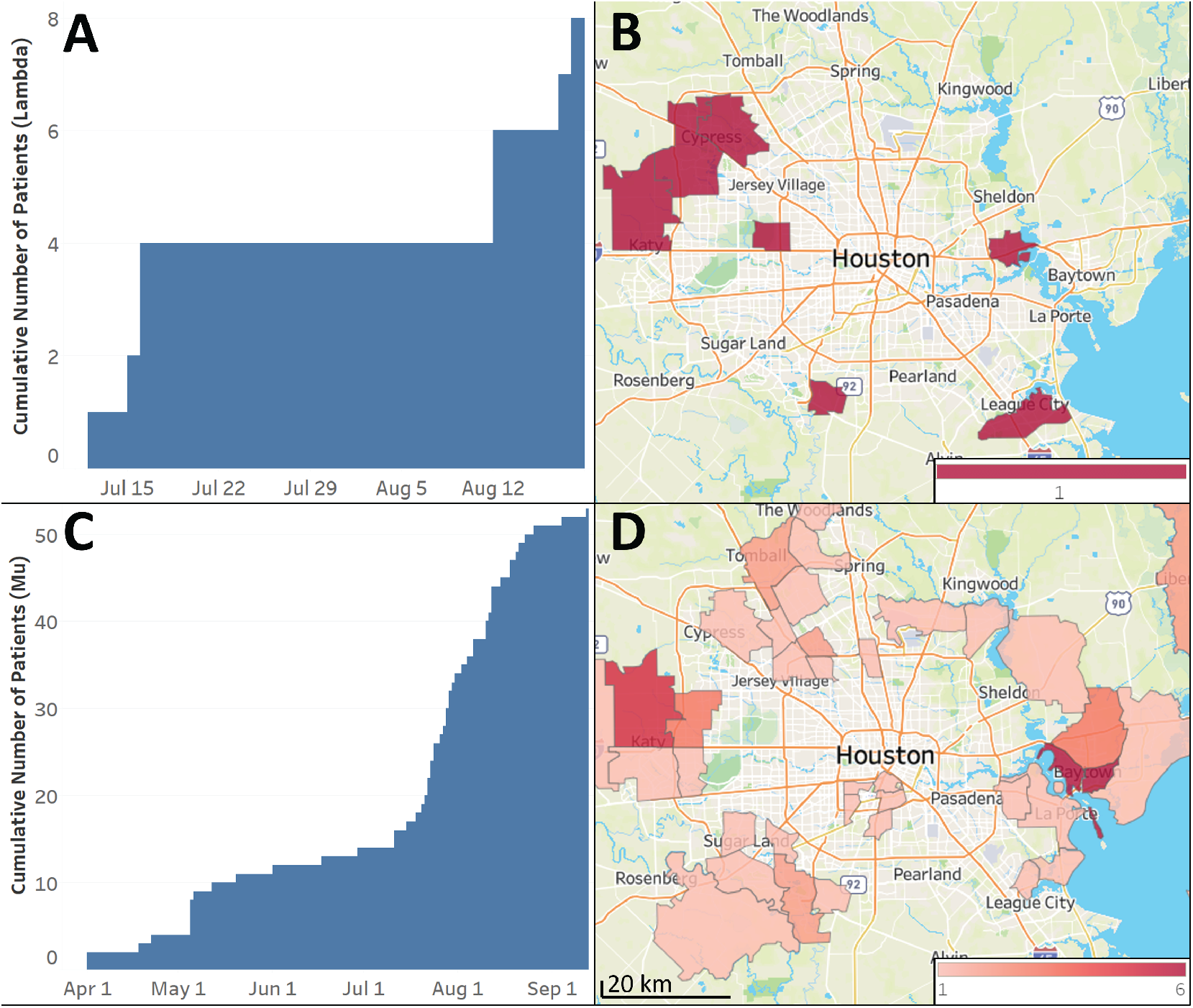
Lambda and Mu variants identified in metropolitan Houston in our patient population. **A:** cumulative increase in the cases caused by Lambda variants identified in the study; **B:** geospatial distribution of these Lambda variants based on the home address zip code for each patient. **C:** cumulative increase in cases caused by the Mu variants identified in the study; **D:** geospatial distribution of the Mu variants based on the home address zip code for each patient. Figures were generated using Tableau version 2020.3.4. (Tableau Software, LLC, Seattle, WA).

## Discussion

In this work, we analyzed SARS-CoV-2 Delta VOCs population genomics and patient characteristics for 16,965 patients, focusing on mid-March 2021 through late September 2021, a time frame in which there was total replacement of the previously dominant Alpha (B.1.1.7) VOC by Delta family VOCs. During this six-month period, a substantial increase in COVID-19 cases occurred in our healthcare system and throughout all of the Houston metropolitan area, virtually all driven by rapid dissemination of the highly contagious Delta family variants. The study was based predominantly on genome sequence analysis of 16,965 SARS-CoV-2 samples from socioeconomically, geographically, and ethnically diverse patients. Several key findings were made, including (i) Delta family VOCs supplanted the Alpha VOC in a relatively short period of time, (ii) regardless of vaccination status, on initial diagnosis, compared to patients infected by other VOCs, patients infected by Delta VOCs had significantly lower Ct values, likely indicating significantly higher viral load in the nasopharynx, (iii) Delta caused significantly more vaccine breakthrough cases than other VOCs, (iv) significantly fewer fully vaccinated individuals required hospitalization, (v) the Lambda and Mu VOIs were identified but were very rare, and they did not increase to substantial levels in the time frame studied.

Our genome sequence data document a rapid increase of Delta variant cases in metropolitan Houston and a corresponding decrease of COVID-19 cases caused by Alpha and other variants, findings similar to epidemiologic trends observed in the UK (https://www.who.int/publications/m/item/weekly-epidemiological-update-on-covid-19---13-july-2021, last accessed August 18, 2021; https://www.ons.gov.uk/peoplepopulationandcommunity/healthandsocialcare/conditions/anddiseases/bulletins/coronaviruscovid19infectionsurveypilot/9july2021, last accessed August 18, 2021). In less than five months, Delta VOCs increased from an initial documented case in our large patient population to cause 99.9% of all cases in our healthcare system. The rapid increase in Delta cases we documented is responsible for a prominent fourth wave of COVID-19 disease in Houston that is ongoing (**Figure 1**). Our findings are consistent with Delta VOCs epidemiologic data reported from other regions of the US.

We found that Delta was significantly more likely to cause vaccine breakthrough cases (**Table 1 and Table 3**). However, importantly, 19.7% of all of our 16,965 COVID-19 cases with genome sequence data occurred in fully vaccinated individuals, but importantly only a small number (n = 896; 5.3%) of these patients required hospitalization. Vaccine breakthrough cases have emerged as an area of great interest, especially so with the increasing percentage of COVID-19 cases caused by Delta variants and the recognition that they are important causes of breakthroughs^31-36^. Although our analysis did not identify a simple relationship between the time elapsed since administration of the second booster vaccination and the date of vaccination breakthrough, this is an important area for continued study. Similarly, we did not study the potential relationship between vaccination breakthrough and waning immunity, but studies of this topic are ongoing.

Some investigators have speculated that the Lambda and Mu variants may become a major concern in future surges. Although in principle this is possible, our data show that neither variant has increased substantially in our metropolitan area during the time frame studied. Thus, our genome analysis of a large set of samples from the first nine months of 2021 does not currently support this speculation, although circumstances may change in the future. Because we are sequencing the genome of the great majority of SARS-CoV-2 causing COVID-19 in our diverse patient population, we are continuously monitoring the growth trajectory of these and other variants in a major metropolitan region in the US. Thus, our ongoing near-real-time sequencing of SARS-CoV-2 genomes responsible for COVID-19 cases in metropolitan Houston provides a facile strategy to assess changes in the virus population composition and molecular evolution in this populous area.

Our study has several limitations that should be noted. Although we sequenced the genomes of SARS-CoV-2 causing 91% of all Houston Methodist COVID-19 cases in the study period, this sample represents only approximately 5% of cases reported in the metropolitan region. We believe it is possible that our patient populations underrepresent certain demographic groups, for example including homeless individuals and pediatric patients. The great majority of the samples sequenced in this study were obtained from symptomatic patients. Thus, it is possible that our sample failed to identify virus genotypes that are preferentially represented in asymptomatic individuals. However, given that at the end of the study Delta represented 99.9% of all cases, we think it unlikely that variants associated with asymptomatic individuals would be substantially different, although this is a formal possibility.

In the aggregate, our data add critical new information to the finding that these three vaccines are highly efficacious in decreasing severe COVID-19 disease, hospitalizations, and deaths^6, 20, 23^. Further, the present study highlights the importance of analyzing SARS-CoV-2 genome data integrated with linked patient metadata and stresses the need to continue to do this in near-real time as the pandemic continues, the virus evolves, and new variants with potentially increased fitness are generated. Analyses of this type are also important in the context of vaccine formulation and long COVID, an increasing health and economic problem globally.

## Data Availability

Genome data used in this study have been deposited to GISAID.

https://www.gisaid.org

## Acknowledgments

We thank Drs. Marc Boom and Dirk Sostman for their ongoing support and Dr. Sasha Pejerrey for editorial contributions and Dr. Heather McConnell for help with figures. The research was supported by the Houston Methodist Academic Institute Infectious Diseases Fund and many generous Houston philanthropists. James J. Davis was funded in whole or in part with Federal funds from the National Institute of Allergy and Infectious Diseases, National Institutes of Health, Department of Health and Human Services, under Contract No. 75N93019C00076. The funders had no role in the design and conduct of the study; collection, management, analysis, and interpretation of the data; preparation, review, or approval of the manuscript; and decision to submit the manuscript for publication.

The findings and conclusions in this article are those of the authors and do not necessarily reflect the views of the U.S. Army.

We declare that we have no conflict of interest.

## Author Contributions

P.A.C., R.J.O., S.W.L., S.S., and J.M.M. had full access to all study data and take responsibility for the integrity of the data and the accuracy of the data analysis; concept and design by J.M.M., P.A.C., R.J.O., and S.W.L; data acquisition, analysis, or interpretation by all authors; drafting of the manuscript by all authors; statistical analysis by P.A.C.; funding obtained by J.M.M. and J.J.D.; overall supervision by J.M.M; P.A.C., R.J.O., and S.W.L. contributed equally and are co-first authors.

## Institutional Review Board statement

This work was approved by the Houston Methodist Research institutional review board (IRB1010-0199).

## Additional Information

Genome data used in this study have been deposited to GISAID.

## Notes

Funding: This project was supported by the Houston Methodist Academic Institute Infectious Diseases Fund; and supported in whole or in part with federal funds from the National Institute of Allergy and Infectious Diseases, National Institutes of Health, Department of Health and Human Services, under Contract No. 75N93019C00076 (J.J.D. and R.O.).

### Competing Interest Statement

The authors have declared no competing interest.

### Author Declarations

This work was approved by the Houston Methodist Research institutional review board (IRB1010-0199).

### Summary of Updates

Revision includes updated SARS-CoV-2 genome sequence data for Houston Methodist cases occurring through August 26, 2021.

